# Optimized Post-Vaccination Strategies and Preventative Measures for SARS-CoV-2

**DOI:** 10.1101/2021.09.17.21263723

**Authors:** Rowland Pettit, Bo Peng, Patrick Yu, Peter G. Matos, Alexander L. Greninger, Julie McCashin, Christopher Ian Amos

## Abstract

**Introduction:** Since March of 2020, over 210 million SARS-CoV-2 cases have been reported and roughly five billion doses of a SARS-CoV-2 vaccine have been delivered. The rise of the more infectious delta variant has recently indicated the value of reinstating previously relaxed non-pharmacological and test-driven preventative measures. These efforts have been met with resistance, due, in part, to a lack of site-specific quantitative evidence which can justify their value. As vaccination rates continue to increase, a gap in knowledge exists regarding appropriate thresholds for escalation and de-escalation of COVID-19 preventative measures.

**Methods:** We conducted a series of simulation experiments, trialing the spread of SARS-CoV-2 virus in a hypothesized working environment that is subject to COVID-19 infections from the surrounding community. We established cohorts of individuals who would, in simulation, work together for a set period of time. With these cohorts, we tested the rates of workplace and community acquired infections based on applied isolation strategies, community infection rates (CIR), scales of testing, non-pharmaceutical interventions, variant predominance’s and testing strategies, vaccination coverages, and vaccination efficacies of the members included. Permuting through each combination of these variables, we estimated expected case counts for 33,462 unique workplace scenarios.

**Results:** When the CIR is 5 new confirmed cases per 100,000 or fewer, and at 50% of the workforce is vaccinated with a 95% efficacious vaccine, then testing daily with an antigen-based or PCR based test in only unvaccinated workers will result in less than one infection through 4,800 person weeks. When the community infection rate per 100,000 persons is less than or equal to 60, and the vaccination coverage of the workforce is 100% with 95% vaccine efficacy then no masking or routine testing + isolation strategies are needed to prevent workplace acquired infections regardless of variant predominance. Identifying and isolating workers with antigen-based SARS-CoV-2 testing methods results in the same or fewer workplace acquired infections than testing with polymerase chain reaction (PCR) methods.

**Conclusions:** Specific scenarios exist in which preventative measures taken to prevent SARS-CoV-2 spread, including masking, and testing plus isolation strategies can safely be relaxed. Further, efficacious testing with quarantine strategies exist for implementation in only unvaccinated cohorts in a workplace. Due to shorter turnaround time, antigen-based testing with lower sensitivity is more effective than PCR testing with higher sensitivities in comparable testing strategies. The general reference interactive heatmap we provide can be used for site specific, immediate, parameter-based case count predictions to inform appropriate institutional policy making.

## Introduction

As of August of 2021, 210 million SARS-CoV-2 cases have been reported worldwide, with over four million corresponding COVID-19 attributable deaths.^1^ Multiple counter measures have be implemented to mitigate resilient viral spread. Such measures have included social distancing mandates^2^, public mask wearing^3^, routine testing plus quarantine strategies^4,5^, and, as of December 2021, mass population vaccination campaigns.^6,7^ As of August 2021, nearly 5 billion doses of a SARS-CoV-2 vaccine have been administered with 37.2 million being administered daily around the globe.^8,9^ This vaccination coverage corresponds to 30% of the world population having received at least one COVID-19 vaccination, and 24% of the world being fully vaccinated.^8^

Occurring in parallel, multiple strains of SARS-CoV-2 have arisen.^10^ The B.1.617.2 (delta) variant is currently the most common variant in the United States and has been since April of 2021.^11^ The delta variant has mutations in the spike protein, and also has demonstrated a higher replication rate, viral loads and transmissibility.^12,13^ Further the widely distributed mRNA vaccines (BNT162b2^14^, mRNA-1273^15^) and adenoviral vaccines (ChAdOx1 nCoV-19^16^) demonstrate attenuated, although still considerable efficacy against the delta and alpha SARS-CoV-2 variants.^17,18^ These variant specific parameters are concerning and have prompted the reinstatement of tighter public health efforts in several areas.^19–22^ As many of these measures, such as social distancing, mask wearing and routine testing + quarantine methods were relaxed following a global decrease in the original SARS-CoV-2 strain^23^, reinstatement has been met with inertia.

A problem exists with providing situation specific evidence for both escalation of counter COVID-19 measures as well as for de-escalation. Public policy measures are muted in efficacy without participant adherence^24^, and current global, national, or regional level guidelines may be viewed as non-specific.^25^ It is not immediately clear to a general audience how small changes in viral parameters can or should direct an organization to scale up or safely wind down their workplace health safety measures.^26–28^ While literature exists and is accumulating on predicting Covid-19 cases nationally^29,30^ or regionally^31–33^, there is little applicable evidence available for reference at the business practice level to inform situation specific decision-making. The viral, community, and workplace parameter thresholds at which a low-risk or a complete COVID-free workplace could be maintained, given changing rates of vaccination adherence and viral variant distributions, remains unattended.

To directly address this question, we conducted a series of microsimulations that simulate the spread of SARS-CoV-2 virus within workplaces that are subject to infections from surrounding communities, using a COVID-19 Outbreak Simulator^34^ that was specifically designed for risk assessment and continuity planning for COVID-19 outbreaks. Our aim was to establish a quantitative reference estimating community and workplace viral spread that would be readily digestible for business organizational use. In our simulations we permute numerous combinations of community infection rates, SARS-CoV-2 variant predominance’s, workplace vaccination coverages, vaccination efficacies, testing with quarantine strategy implementations, SARS-CoV-2 test efficacies, and routine indoor mask wearing, and observe how expected case counts change for cohorts of individuals working together. We identify appropriate circumstances where masking and testing strategies may be discontinued or should be implemented. Further, we present the impact of vaccination coverage in the workplace on the efficacy of preventative SARS-CoV-2 testing strategies. We finally introduce generalizable reference heatmaps which can allow for quick identification of appropriate mitigation strategies for an organization given site-specific details. We hope that our presentation of these simulation findings will allow for quantifiable and transparent policy decision-making that can respond to shifting viral, organizational, and community specified parameters.

## Methods

### COVID-19 Outbreak Simulator

We modified and applied the COVID-19 Outbreak population-based simulator^34^ which simulates the spread of SARS-CoV-2 virus in dynamic and heterogeneous populations. We estimated relevant SARS-CoV-2 parameters from the clinical literature for viral transmission dynamics and used the simulator to observe parameter specific viral outbreak outcomes in feed-forward time simulations. We modified the model to also simulate the impact of vaccination on viral transmission in generated scenarios. We modeled SARS-CoV-2 testing using both RT-PCR and Antigen tests^5^ and implemented a plugin for COVID-19 vaccination with varying vaccination coverage and vaccination efficacy. Additionally, we introduced and observed the effect of varied sizes and durations of work groups gathering, testing with quarantine strategies, community infection rates, scales of testing, non-pharmaceutical interventions, variant predominance’s and testing strategies on expected case counts. In total we set up 33,462 scenarios, to provide quantitative estimates of workplace and community associated infections expected for each.

### Simulated Workplace Environment

We simulated a generalizable workplace environment in which individuals residing in a community come into contact in a shared location daily for common tasks. Viral SARS-CoV-2 transmission occurs during interactions in the community and within interactions in the workplace. More specifically, team members are exposed to the community and are subject to a “community infection rate.” This is a location-dependent parameter that can be determined by reference to current and projected rates of infection in specific areas. An infected individual could infect one or more team members within the workplace. The probability of an infected individual infecting others (reproduction number R0), or transmission rate, is reduced in our simulations by the effect of non-pharmaceutical interventions (NPI), testing and vaccination. The simulated environment we created assumes members work five days per week with two days off. We modeled two general work team scenarios. The first included an office of 60 members interacting together for a 8-week period (480 person-weeks). A second set of simulations included an office of 200 individuals working together for 24 weeks (4,800 person-weeks).

### Virus Specific Parameters

Community infection rate (CIR) is the actual community rate of infection and is used to model the probability that anyone in the community will be infected per day prior to coming to the workplace. For these simulations we have used community infection rates of 0.5,1, 2.5, 5, 10, 15, 30, 60, 90, 120 and 150 infections per 100 thousand people per day. We assumed that 30% of all infected individuals will remain asymptomatic according to model recommendations from the United States Centers for Disease Control and Prevention (CDC). We assigned a variant specific random reproduction number to each infected individual, which will determine, on average, the number of individuals each will infect during their infection. For simulations representing the original SARS-CoV-2 strain, reproduction numbers were drawn from random distributions with mean of 2.5 for symptomatic cases, and of 0.5 for asymptomatic carriers. For simulations of the alpha (B.1.1.7) strain, the randomly assigned distribution mean was 3 for symptomatic cases and 2.1 for asymptomatic carriers. For simulations of the delta (B.1.617.2) the R0 distribution mean was 6.5 with a 95% confidence interval from 5 to 8 for symptomatic carriers and a mean of 4.5 for asymptomatic carriers. We assumed that the transmissibility of asymptomatic carriers are 70% of those of symptomatic cases for the alpha and delta variants according to CDC recommendation.

### Non-Pharmacological Interventions

We used a “distancing factor” to model the effect of non-pharmaceutical interventions (NPI) measures including mask wearing and physical distancing. This distancing factor changes the reproduction number of individuals during simulation but does not affect individual their viral loads. We modeled scenarios with no mask wearing, with continuous mask wearing (decreased R0) and with no mask wearing and overcrowding throughout the workday. To represent masking, a factor of 0.6^35^ was multiplied to an individual’s overall transmissibility while wearing a mask. A factor of 1 represented no NPI, and a factor of 1.2 was multiplied to overall transmissibility to represent overcrowding and no masking. An overcrowding scenario was included to represent industries such as movie productions during which team members often stay in close contact for extended period of time such as when filming indoor scenes.

### Testing Strategies

In our simulations we included implementation of routine COVID-19 testing strategies. For these simulations we include testing team-members for the presence of SARS-CoV-2 with either a polymerase chain reaction (PCR) or an antigen-based test. To accurately present the efficacy of testing we simulated testing strategies using two SARS-CoV-2 tests, a RT-PCR test with a clinical sensitivity of 90% and an antigen test method with a clinical sensitivity of 80%, which corresponds to two commonly used tests observed in the literature^5^ In **Table 1**, we present the clinical sensitivities, specificities, and turnaround times for simulated testing scenarios. Also included in Table 1 are the frequency with which these tests were conducted in our simulated scenarios, which includes not testing (None), testing once a week (on Monday – M), testing three times a week (MWF), to testing daily during the work week (MTWTF). We start each week from Saturday and start testing at the beginning of the third day (Monday). We define clinical sensitivity as the sensitivity of a test when it is applied to a population with people at different stages of infection in a clinical setting, mostly determined by the limit of detection of the test and the viral load of the sample at the time of testing. Specificity is the probably that a test correctly generates a negative result when an individual was not infected. Turnaround time is the time required for the test results to become available. In our model, infected team members return to work after samples are collected and could infect others before the test results become available. To simulate testing with quarantine in our simulated workplace environment, team members were isolated for 10 days if they showed any COVID-19 symptoms or tested positive by either a RT-PCR based test or an Antigen test. We differentiated between true and false positive tests and only isolated truly infected team members, because isolating negative individuals will not affect future transmissions. Asymptomatic carriers could infect others and remain undetected. Infected team members who recovered from an infection could still be infectious after isolation but had markedly decreased transmission probability (1%). We assume that workplaces will continue with the same number of members, representing temporary replacements, even if one or two members are in isolation. We simulated testing strategies applied to all team members, and additionally applied exclusively toward team members who were not vaccinated.

**Table 1.**
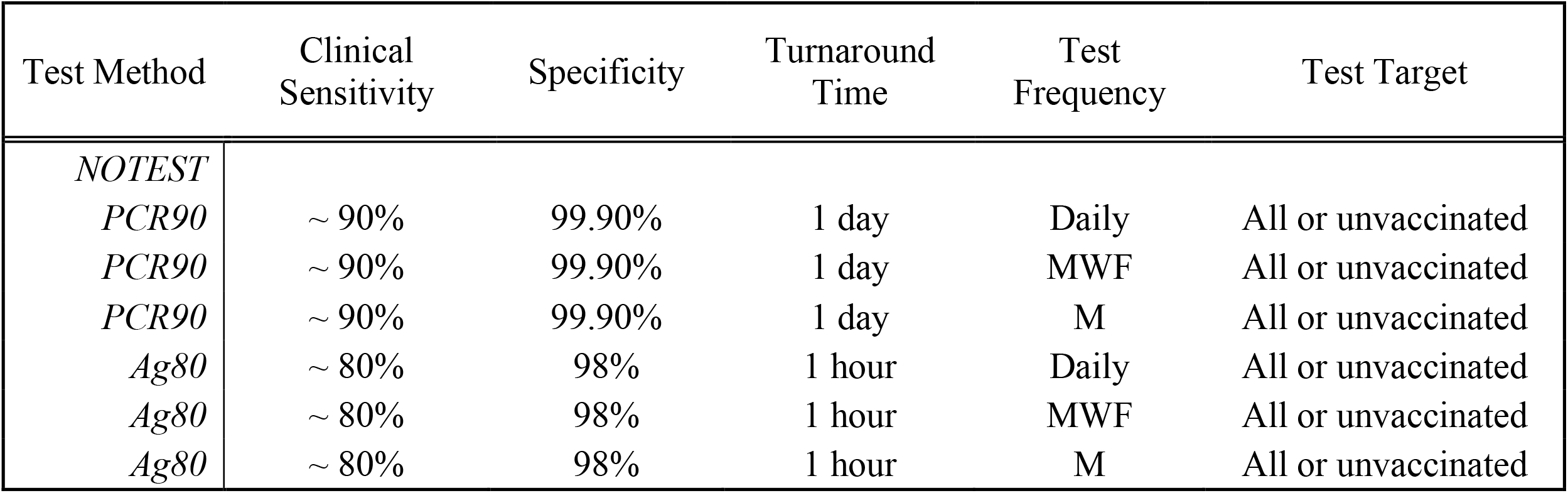
Sensitivity and Specificity of simulated SARS-CoV-2 testing methods

### Vaccination Efficacy and Coverage

We varied the parameters related to vaccination of team members in our simulations. To model vaccination, we jointly modulated the probability of infection (immunity to infection), and the probability of transmission (infectivity after vaccination) in vaccinated individuals. We modeled vaccines with 95% efficacy, 70% efficacy, and 50% efficacy. These probabilities represent the post-vaccination immunity to infection, or the probability that a vaccinated individual is immune to an infection event. We modulated vaccinated individuals to have a decreased infectivity after vaccination to 5% of their infectivity baseline. This was done to represent their diminished viral load that could cause an infection to others. Specifically, if an individual (unvaccinated) had a viral load that would on average infect three people during an infection, then a Covid-19 positive vaccinated individual would on average infect 3 persons * 0.05, or 0.15 people after vaccination. Additionally, if an individual is vaccinated with a vaccine with 70% efficacy, the individual would be able to avoid an infection event at the probability of 0.70. Finally, in a workforce population, we included in our simulations a ‘level of coverage’ parameter to observe the effects of varying the percentage of vaccinated individuals in the workplace. We allowed the ‘coverage’ parameter to range from 0% (no vaccinated team members), 25%, 50%, 75%, or 100%. For simplicity, we assume full vaccination according to CDC definition and do not consider variation of efficacy and the increase (*e*.*g*., between first and second shots) and loss of immunity (*e*.*g*., out of protection period). **Table 2** delineates all the permutations of vaccine interventions we included in these simulations.

**Table 2.**
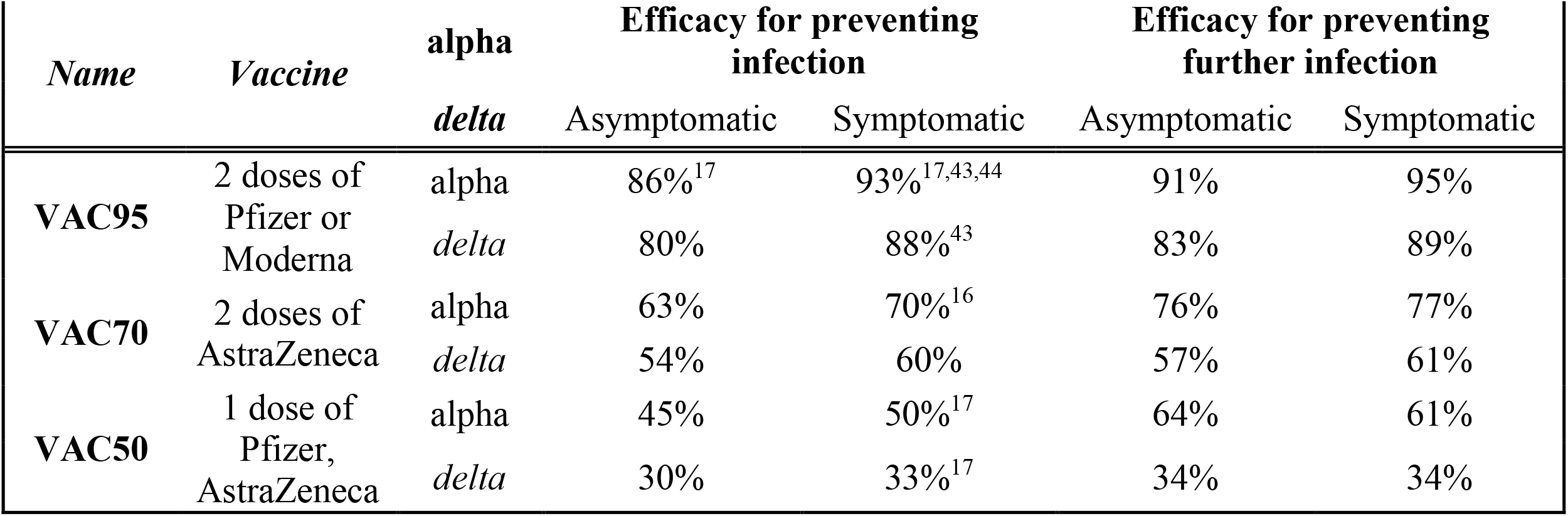
Permutations of Vaccination Efficacy in Workplace Interventions

### Outcomes Measurement

During the course of the simulations, we measured and report the number of community acquired infections (CAI), the number of workplace acquired infections (WAI), and the total number of acquired infections (TAI, of CAI + WAI). We selected WAI to be our main output metric. We measure and report WAI over 480 person-weeks (PW) and 4,800 PW, representing our 60-persons followed for 8 weeks and 200 persons followed 24 weeks cohorts respectively. To determine efficacy of intervention, we set an acceptable range threshold for WAI to be less than one. For ease of generalization of results, we also report a scaled down WAI, CAI and TAI to per-person per-week metric outcomes. Finally, we also track and report the total number of symptomatic workforce members, number of infections detected, and the number of false positive tests expected per scenario. All scenario outcome metrics may be viewed in **Table 3** (supplement only). The simulation package is publicly available at https://ictr.github.io/covid19-outbreak-simulator/.

## Results

Using the COVID-19 Outbreak Simulator we simulated 33,462 scenarios with various community infection rates, vaccine types and coverages, non-pharmaceutical interventions (NPI), and testing strategies. In **Table 3**, we report for each of these 33,462 combinations of input variable parameters and output metrics. Summary heatmaps of our findings regarding simulated total cases per scenario are presented as **Figures 1-4**. Each figure presents a multi stratified heatmap which annotates infections expected after each simulation based on community infection rate, vaccination coverage percentage, vaccination efficacy, and SARS-Cov-2 strain, which is stratified by various ‘testing + quarantine’ strategies. We repeat this analysis twelve times, iteratively testing the additional effect of testing + quarantine strategies applied to all employees versus to only unvaccinated employees, and with or without concomitant mask wearing while in the office place, and for two working group size + duration combinations. **Figure 1** presents WAI through 480 PW without masking with testing for all employees, while **Figure 2** presents with masking and testing for all employees. **Figure 3** presents TAI, without masking, through 480 PW. **Figure 4** presents WAI through 4,800 PW without masking with testing for all employees, while **Figure 5** presents with masking and testing for all employees. **Figure 6** presents TAI through 4,800 PW. All permutations of our results are displayed online, with push button toggle features, for public presentation https://public.tableau.com/app/profile/rowland.pettit/viz/COVID-19Heatmaps/COVID-19Simulations.

**Figure 1.**
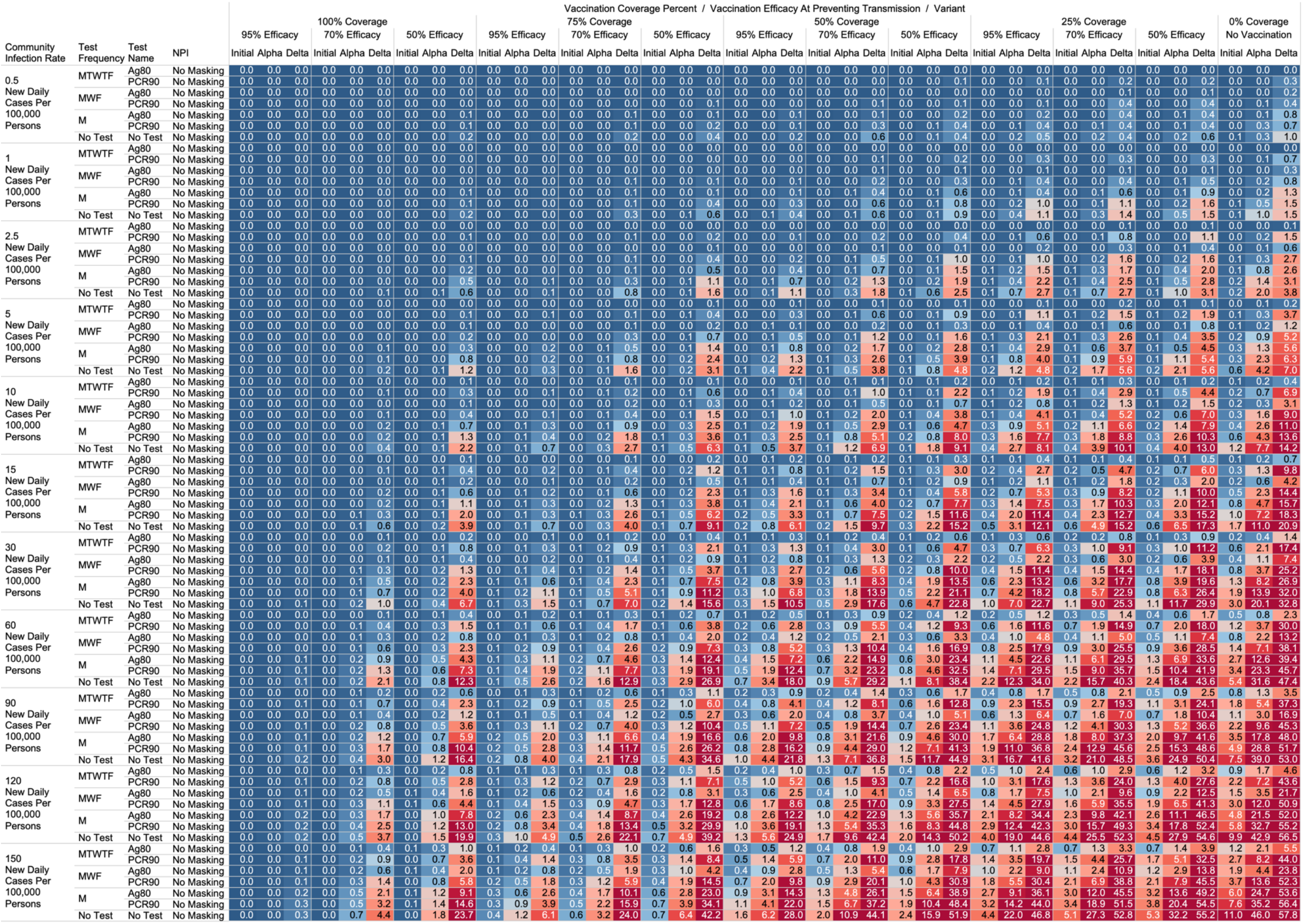
Heatmap of simulated workplace acquired infections through 480 person weeks with testing performed in all workplace members and with no masking intervention.

**Figure 2.**
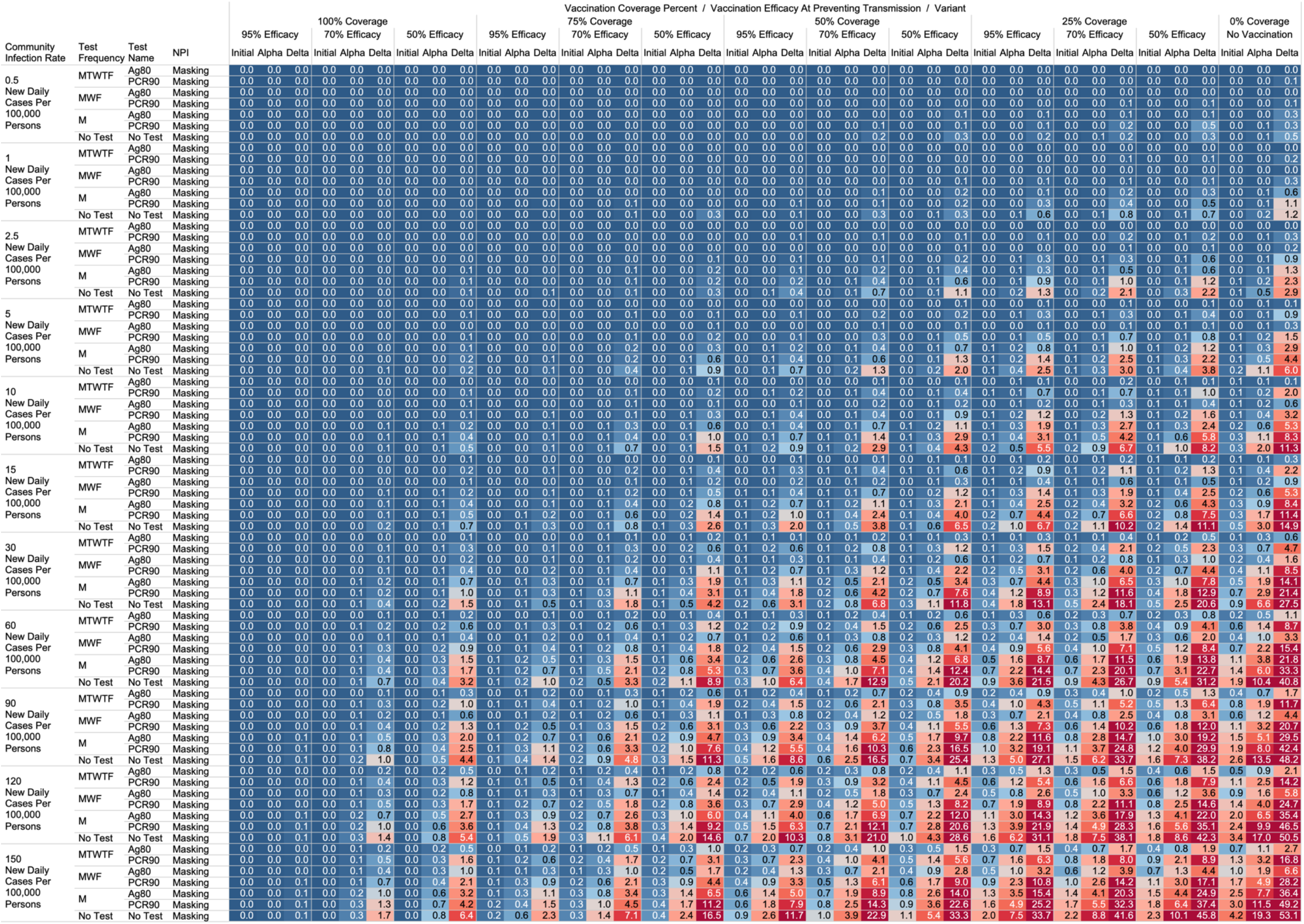
Heatmap of simulated workplace acquired infections through 480 person weeks with testing performed in all workplace members with consistent mask wearing.

**Figure 3.**
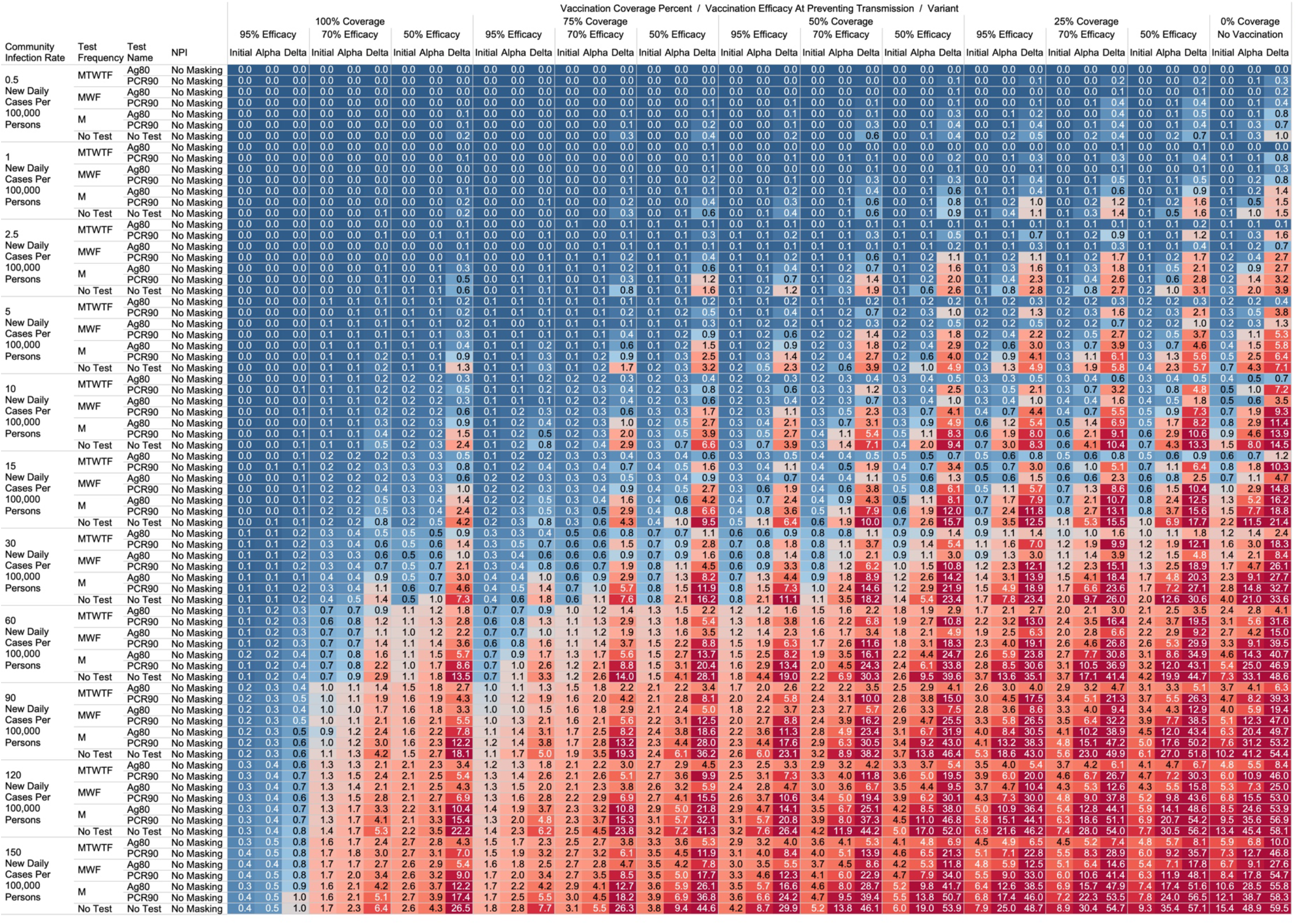
Heatmap of simulated total acquired infections (CAI + WAI) through 480 person weeks with testing performed in all workplace members and with no masking intervention.

**Figure 4.**
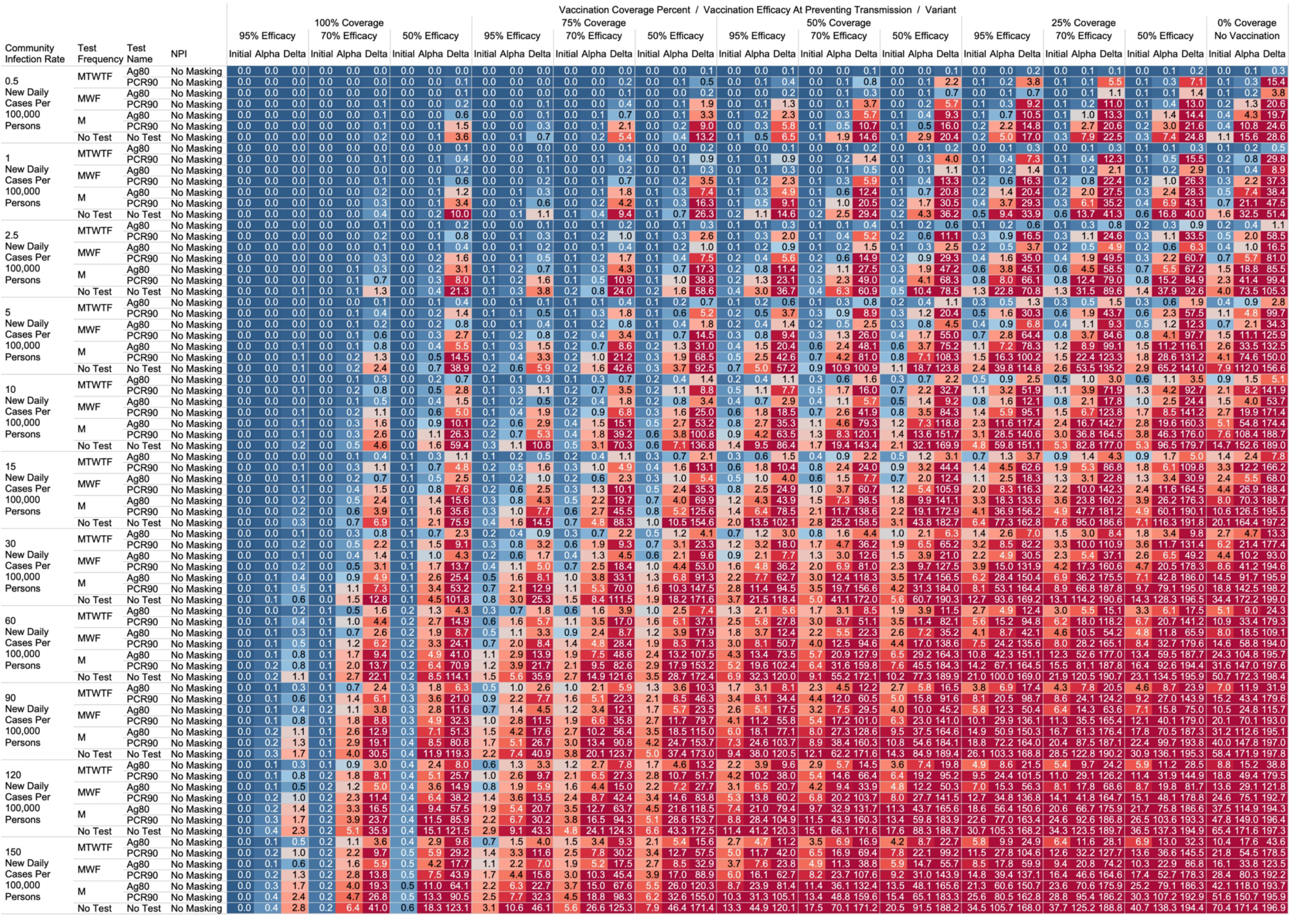
Heatmap of simulated workplace acquired infections through 4,800 person weeks with testing performed in all workplace members and with no masking intervention.

**Figure 5.**
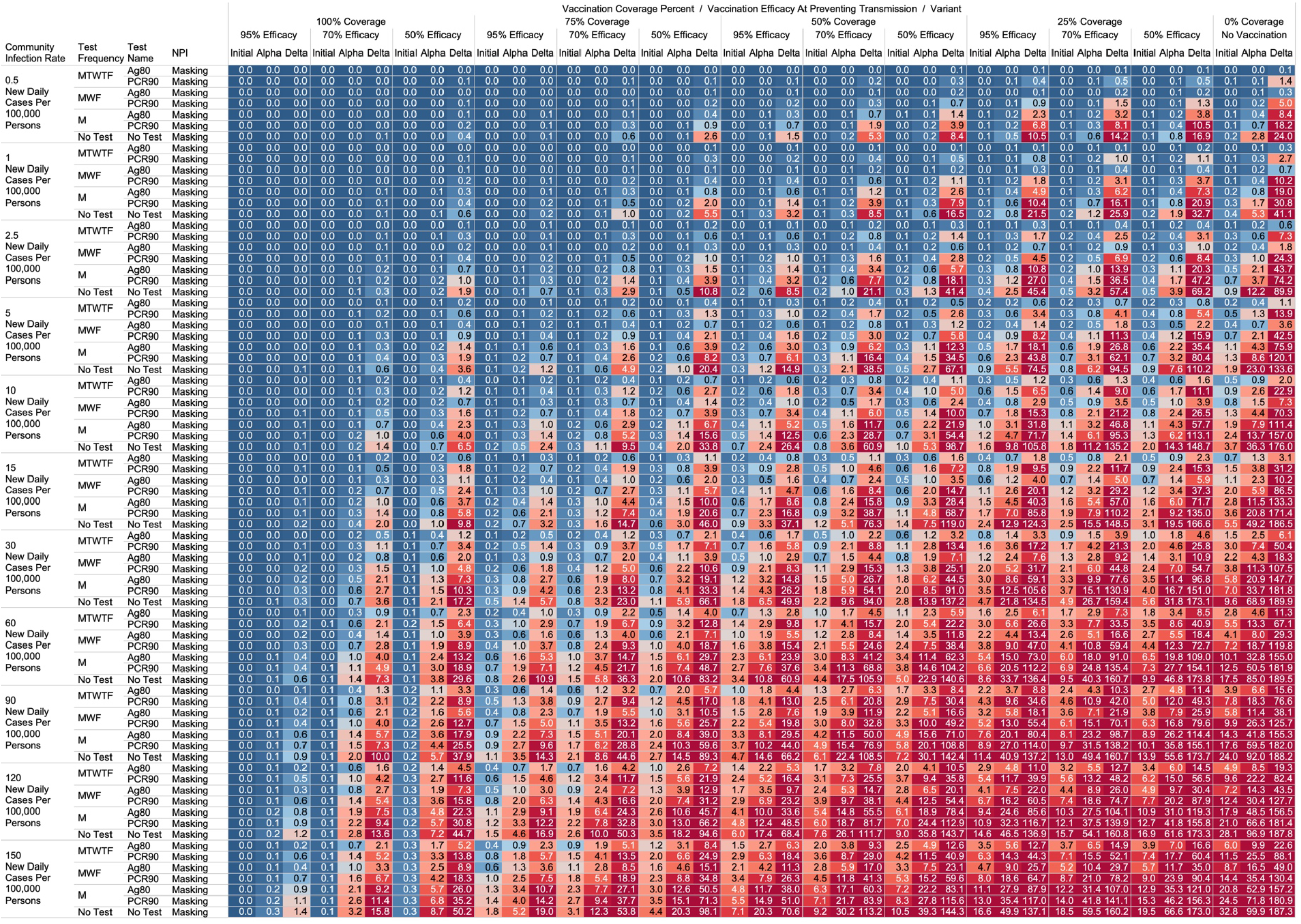
Heatmap of simulated workplace acquired infections through 4,800 person weeks with testing performed in all workplace members with consistent mask wearing.

**Figure 6.**
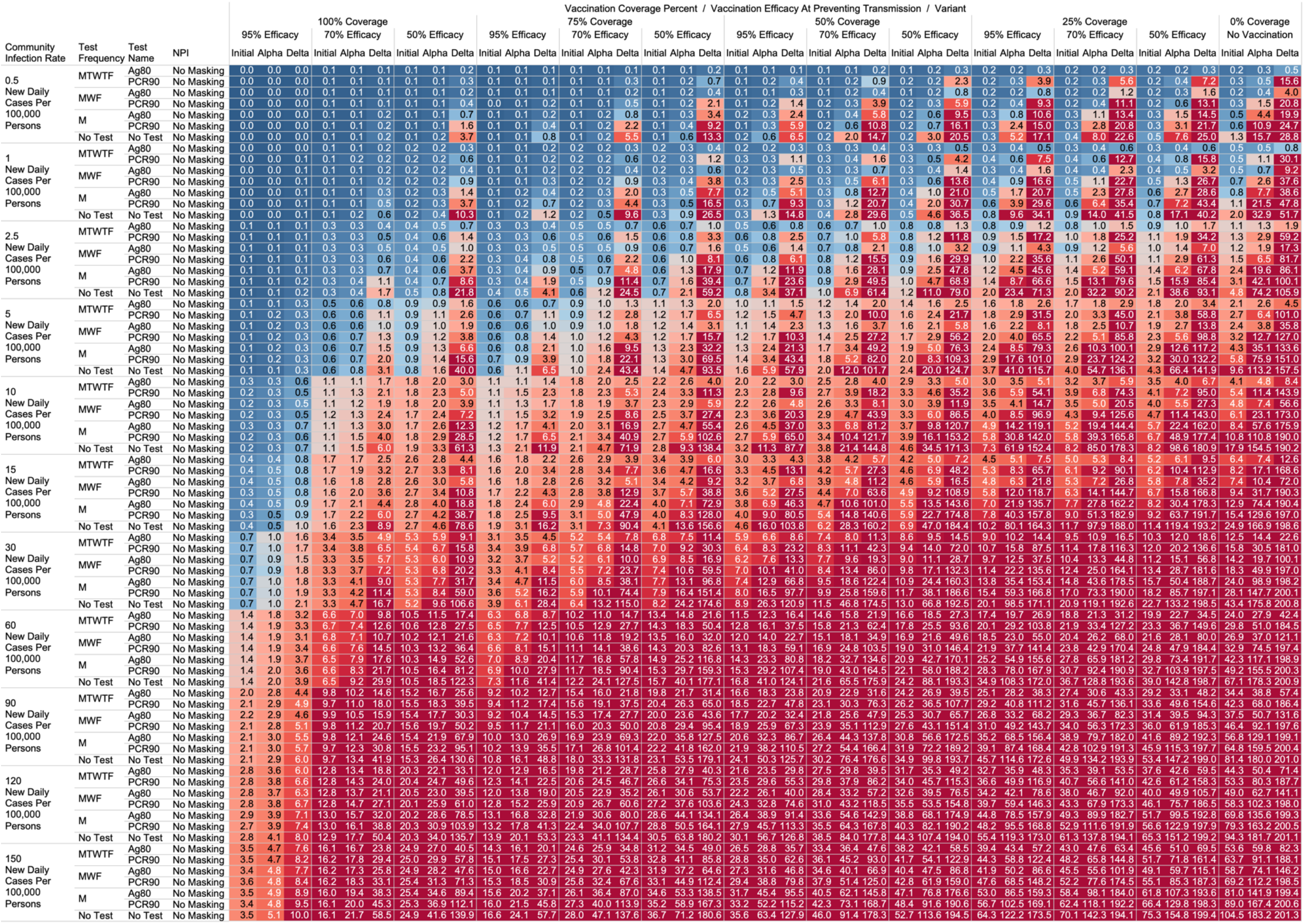
Heatmap of simulated total acquired infections (CAI + WAI) through 4,800 person weeks with testing performed in all workplace members and with no masking intervention.

### No Masking Criteria

With a threshold of less than or equal to one workplace aquired infection per cohort specific person weeks, we found some conditions in which no masking is necessary. If the community infection rate (CIR) per 100,000 persons is less than or equal to 150, and the vaccination coverage of the workforce is 100% with 95% vaccine efficacy then no masking or routine testing + quarantine strategies are needed to achieve less than one workplace acquired infection WAI in most scenarios. Specifically, when following either the 60 persons by 8-week work group (480 PW) or the 200 persons by 24-week work group (4800 PW) the expected WAI in settings dominated by the original SARS-Cov-2 (1°) strain is 0.000 and 0.004 respectively. In settings dominated by the alpha (α) strains, expected WAI is 0.037 and 0.446 for 480 PW and 4800 PW respectively. In preventing the delta (Δ) strain, the no test no mask strategy was effective after 480 PW (WAI = 0.275), but not through 4,800 PW (WAI = 2.81). The minimum CIR which yielded less than one WAI with no testing and no masking against the delta variant through 4,800 PW was less than or equal to 30 new daily cases per 100,000 persons (WAI = 0.62). As no testing is necessary, there will be no false positive tests and no expected symptomatic workforce members.

For less efficacious vaccines, at efficacies of 70%, testing and masking measures may be still completely relaxed regardless of variant predominance if the vaccination coverage is 100% and the community infection rate is 1 or less (WAI expected through 480 PW 1° = 0.000, α = 0.004, Δ = 0.039; 4,800 PW 1° = 0.001, α = 0.043, Δ = 0.431). No masking and no testing resulted in less than one case in several other specific settings, although not for all variants. Obtaining 75% workplace vaccination coverage with a 95% efficacy vaccine would result in less than one WAI as long as CIR is 10 or fewer through 480 PW but not the 4,800 PW (480 PW 1° = 0.027, α = 0.103, Δ = 0.670; 4,800 PW 1° = 0.302, α = 1.067, Δ =10.815). These trends, and other effective no testing, no masking combinations can be appreciated visually in Figures 1 and 3, given individual site-specific parameters.

### Optimal Testing Strategies

Overall, more frequent testing plus isolation strategies decreased the probability of infection. For example, when routinely testing all employees, without concurrent masking, the daily Monday-Tuesday-Wednesday-Thursday-Friday (MTWTF) testing strategy resulted in fewer WAI than the MWF, which was superior to a Monday only method, which was better than no testing. As an example, looking at utilizing a PCR test with a 90% sensitivity or true positive rate and a community infection rate of 15 new daily cases of the original variant per 100,000, and no vaccination coverage, the WAI expected with MTWTF testing is 0.34 after 480 PW, while for MWF 0.48, for M only 0.99, and 1.69 for no testing + quarantine interventions. This trend held whether for antigen versus polymerase chain reaction (PCR) testing, with or without masking, and for each viral strain.

Further, we report an observed trend that, comparing within a testing frequency, testing and isolation with an antigen-based testing method resulted in the same or fewer WAI than testing with polymerase chain reaction (PCR) methods. Despite the decreased sensitivity, we report that with no masking and no vaccination coverage, a community infection rate of 15 for the original strain, and a MWF testing strategy, the WAI when testing with an 80% sensitive antigen test is 0.22 per 480 person weeks but 0.48 with a 90% sensitive PCR test. In the same setting except a CIR of 15 for the alpha variant the expected infections are 0.59 for the 80% sensitive antigen test, and 2.33 for the 90% PCR. Finally, in the same scenario with CIR of 15 for the delta variant with MWF testing, expected WAI are 4.22 with 80% sensitive antigen test and 14.35 with 90% sensitive PCR test. This trend holds for other testing frequencies and community infection rate scenarios.

### Vaccination Status Based Testing

We report on the efficacy of only routinely testing and isolating non-vaccinated individuals in the workforce. When the community acquired infection rate is 5 new confirmed cases per 100,000 or fewer, and at 50% of the workforce is vaccinated with a 95% efficacious vaccine, then testing daily with an antigen-based test in only unvaccinated workers will result in less than one WAI (480 PW 1° = 0.010, α = 0.025, Δ = 0.05; 4,800 PW 1° = 0.123, α = 0.264, Δ =0.872) without the need for workplace masking. Similarly, a MTWTF testing regimen in only unvaccinated workers with a 80% sensitive antigen test yielded one WAI independent of variant predominance in situations of no vaccination coverage (480 PW 1° = 0.017, α = 0.041, Δ = 0.111; 4,800 PW 1° = 0.221, α = 0.441, Δ =1.416) when the CIR is 2.5 or fewer.

One day a week testing regimens (Monday only) with an 80% sensitive test only in unvaccinated populations results in less than one case, and are superior to no test strategies, in specific scenarios. These include when the CIR is less or equal to 2.5 and 55% of workers are vaccinated with a 95% efficacious vaccine (480 PW WAI 1° =0.02, α = 0.06, Δ =0.42). Several other unvaccinated only testing strategies with different test types were observed to result in less than one infection per scenario at higher community infection rates than mentioned as vaccine coverage, efficacy and NPI interventions were introduced. These specific findings may be appreciated in **Figures 1-4**.

### Total Acquired Infections

With a threshold of less than or equal to one total infection per cohort, no testing or masking was needed when CIR was 15 or less with 100% coverage of a 95% efficacy vaccine occurred in the cohort (TAI expected after 480 PW 1° = 0.036, α = 0.051, Δ = 0.126; 4,800 PW 1° = 0.351, α = 0.526, Δ =1.044). With no vaccination coverage, less than one TAI was only observed when CIR is less than or equal to 1, masking was implemented, and a MWF or more frequent testing strategy with an 80% sensitive antigen test was used (TAI expected 480 PW 1° = 0.044, α = 0.034 Δ = 0.055; 4,800 PW 1° = 0.376, α = 0.383, Δ =0.6125). At least one CAI can be expected at 4,800 PW independent of masking, testing strategies or vaccination coverages when CIR is 60 or greater.

## Discussion

We used a population-based simulator that imitates the spread of SARS-CoV-2 virus in a dynamic and heterogeneous population, to demonstrate the impact of testing, vaccination rates, variant representation, and community infection rates to provide a quantitative analysis of workplace associated infections. We estimated relevant SARS-CoV-2 parameters from the clinical literature for viral transmission dynamics and used the simulator to observe parameter specific viral outbreak outcomes in feed-forward time simulations. We included vaccination coverage and vaccination efficacy as new model parameters to provide estimates of effectiveness of SARS-CoV-2 outbreak mitigation measures. In practice our 95% vaccine efficacy can be used to represent individuals who have received two doses of the Pfizer or Moderna Vaccine, 70% efficacy would be for individuals who received two doses of AstraZeneca, and 50% efficacy would be individuals who have received one dose of Pfizer or AstraZeneca vaccines against the delta B.1.617.2 variant.^17^ However, reports on efficacy of vaccines against specific variant are incomplete and often inconsistent. For example, more recent reports showed that the efficacy of the Pfizer vaccine is lower than initially reported (42%), so results from VAC50 could be used for Pfizer in communities with infections dominated with the delta variant.

Our 33,462 simulations identified pertinent thresholds. Several scenarios exist in which preventative measures taken to prevent SARS-CoV-2 spread, including masking, and testing plus quarantine strategies can safely be relaxed. These thresholds were found to vary substantially based on local variant predominance. Where the alpha or original variant predominate the CIR thresholds, the limits at which no masking or routine testing would be necessary were often more than double than what would be sufficient to have no cases of the delta variant. For example, with 100% coverage of a 95% effective vaccine in a workplace, the CIR at which no testing or masking would be required was 150+ for the original strain but 30 for the delta variant through 4800 PW. There are specific plausible scenarios where neither masking nor testing are needed to operate safely in a workplace, contingent on CIR, vaccination coverage, and vaccination efficacy metrics. We view that the quantitative and granular estimates we present can serve to benchmark disease mitigation performance that will be needed to maintain acceptable risks in workplace environments.

We also quantify the efficacy of testing + isolation strategies in the context of a sliding scale of increasing vaccination coverage and varied vaccine efficacies. The main finding regarding testing showed that using antigen testing with 80% sensitivity three times per week compared to PCR testing with a 90% sensitivity three times per week regimen gives the same or better results. Despite the lower sensitivities of the antigen testing methods, antigen testing yields similar or superior results to PCR testing likely due to quicker turnaround. Finally, we report the specific finding that in workplaces implementing routine mask wearing, daily testing of only unvaccinated members is effective to prevent 1 workplace acquired infection per 480 PW when the CIR is 30 or fewer. Similarly, less than one WAI is predicted through 4800 when the CIR is 2.5 or fewer, independent of workplace vaccination coverage and or local strain predominance.

While we view these individual findings to be useful, we feel a broader and more immediate utility of this work may be a realized through publication of these expected infection heatmaps to a public audience. Figures 1-4, and accompanying online interactive dashboard https://public.tableau.com/app/profile/rowland.pettit/viz/COVID-19Heatmaps/COVID-19Simulations, afford realistic and quantitative projections of viral spread based on relevant parameters. These estimates clearly present exactly how changes in relevant parameters will lead to acceptable or unacceptable case count projected risks. By presenting our results in this manner, we hope that workplace clinical and administrative teams could precisely set their policies, accounting for parameter changes in their workplace environment and community as they occur immediately. For example, one can identify their community infection rate, their employee vaccination coverage and vaccine type efficacy and then benchmark what they feel is an acceptable level of risk for their employees. A specific testing strategy, occurring in all employees or just in vaccinated individuals, with a certain test type/sensitivity, with or without the use of masking can be identified to meet specific workplace needs. As CIR or other parameters shift, our aim is for such heatmaps to allow for quantitative justification of increasing or relaxing SARS-CoV-2 protocols optimally.

We view this simulation and dissemination of information to be unique, and a complement to resources already available. Currently several national level and state/regional level dashboards exist for assessing global risk. A similar yet non-overlapping concept has been published by Chhatwal et al. as a preprint^33^ whose team hosts a COVID-19 simulator application. This application provides national and regional analysis which allows for relevant viral transmission parameters to be modulated and intervention strategies to be implemented for set durations, and in combination. While useful, we view our model fills a gap not addressed with their model, or other models we have identified, in that it provides quantitative estimates at the institutional level, not as the state level. Estimates of workplace infections can be made per establishment based on highly local and company specific parameter input.

Our study has some limitations. We describe the output of a simulation of two cohorts, 60 individuals commuting to a common workplace for a 80-week period (480 PW) and 200 workers meeting together for 24 weeks (4,800 PW). Certainly, individual workplace environments vary in size ranging from very small groups to very large groups. Interestingly the per person per week simulated infections were highly similar between our 480 PW and 4,800 PW simulated workgroups, indicating our modelling study can be helpful in providing an evidence-informed projection to medium to large workgroups. However, our results do not represent what may happen to small environments due to the well observed phenomenon that workplace outbreaks are often started by super spreader events rather than the steady continued transmission of one infected member to another.^36^ Additionally, more nuanced dynamics exist in which the probability of interaction for some team members may be less than for others, such as when simulating an administrative office versus a warehouse operations cohort all in the same workplace but not as frequently interacting.

We also assume that none of the team members are infected at the beginning of simulations. In practice, this reflects that a negative PCR test is required before members can join the workplace to ensure an uninfected team. We start the simulations from a Saturday so team members will be subject to two days of community infection before starting to work (and probably get tested) on Monday. Our recommendations have factored in these conservative assumptions; however, any implementation of a revised testing strategy should be informed by prospective monitoring of workplace-associated infection rates. We use the CIR or the actual community rate of infection as a parameter to model the probability that anyone in the community will be infected per day prior to coming to the workplace. This number is usually greater than the reported “confirmed” daily test number of cases. The actual number will differ from the confirmed numbers to varying degrees due to reporting delays and the sufficiency of testing^37^. CIR reflects an average infection rate in a region and will vary among people with different occupations, ethnicity, living conditions, educational levels, etc. For many workplace teams, members may have a lower infection rate than the public due to their observance of the disease mitigation protocols.

In our simulations we assumed that 40% of all infected individuals will remain asymptomatic for the original variant and 30% for alpha and delta variants.^38^ This and other viral transmission parameters could be updated as further information is published on each variant. We note that the sensitivity and specificity of SARS-CoV-2 tests will vary by manufacturer, and by region, and a wide range of sensitivities and specificities have been reported from the literature. We modeled a PCR test and an Antigen test with varying sensitivities. We consider the PCR90 as a representative RT-PCR test performed by a typical US clinic^39^; Ag80 as an example for high performance Antigen test such as Abbott BinaxNOW.^40^ However, sensitivity of Antigen tests vary, especially when they are applied to samples with low viral loads. For example, the Innova SARS-CoV-2 antigen rapid lateral flow test that is widely used in England has high sensitivity on samples with higher viral load (91% for samples with Ct < 18.3, 69% for samples with Ct < 24.4) and poor sensitivity otherwise (9.7% for samples with Ct < 24.4).^41^ Our model for antigen test matches the sensitivity of this test for moderate to high viral loads as we generally expect for carriers with the delta variant, but may not represent real-world scenarios for other antigen tests. In a low prevalence situation, both PCR and antigen testing can yield a substantial number of false positive results.

We constrained vaccine coverage in the workplace to be 0 (no vaccination), 25%, 50%, 75%, or 100%. For simplicity, we assume full vaccination according to CDC definition and do not consider variation of efficacy and the increase (*e*.*g*., between first and second shots) and loss of immunity (*e*.*g*., out of protection period). Finally, The model does not account for the impact of quarantining of close contacts to the index case which would occur in the real world in accordance with public health guidance.^42^ The impact of quarantining contacts would be further reduce workplace acquired infections. It would be difficult to model quarantining as the number of contacts would be dependent on the workplace interaction behavior of the index case as well as the ability to successfully identify all close contacts.

With these limitations in mind, we feel we present an evidence based, realistic assumption driven simulation study of SARS-CoV-2 outbreak in a variety of settings. From these analyses we report specific findings including applicable scenarios safely supporting no masking + no testing policies, antigen testing superiority, and unvaccinated only testing strategies. We also present our full heatmaps of expected outbreak outcomes dependent on community infection rate, vaccination coverage, vaccination efficacy, testing strategies, and population masking. These can be utilized by employers and policy makers to predict infection expectations, allowing for data-driven and community specific SARS-CoV-2 preventative strategies. Future investigations will be warranted, and we therefore make all of our algorithms and approaches readily available. As newer strains emerge, updating these heatmaps to reflect the continuing mix of viral strains, with strain specific reproduction numbers should be pursued. Further, expanding these heatmaps to more broadly represent viral transmission outside of a workplace but on a macro city or state level is also justified. For future investigations we invite readers to precisely model their own environment using the https://ictr.github.io/covid19-outbreak-simulator/.^34^ We have provided the baseline parameters to reproduce our work, and several site-specific tutorials exist for modifying our code for site specific constraints.

## Data Availability

The simulation package, which was used to generate our simulation data is publicly available at https://ictr.github.io/covid19-outbreak-simulator/. All simulation results are provided in supplementary Table 3.

## Acknowledgements

The authors would like to thank the Baylor College of Medicine Medical Scientist M.D./Ph.D. Training Program for their support (RWP). The authors would like to thank BRASS: Baylor Research Advocates for Student Scientists for their support (RWP).

